# Technical Optimization of SyntheticMR for the Head and Neck on a 3T MR-Simulator and 1.5T MR-Linac: A Prospective R-IDEAL Stage 2a Technology Innovation Report

**DOI:** 10.1101/2025.04.08.25325491

**Authors:** Lucas McCullum, Samuel L. Mulder, Natalie A. West, Hayden Scott, Diana Carrasco Rojas, Zayne Belal, Cem Dede, Warren Floyd, Alaa Mohamed Shawky Ali, Abdallah Sherif Radwan Mohamed, Alex Dresner, Ergys Subashi, Dan Ma, R. Jason Stafford, Ken-Pin Hwang, Clifton D. Fuller

## Abstract

**Objective:** The purpose of this study was to optimize the technical tradeoffs associated with integrating the quantitative maps available from SyntheticMR into the head and neck adaptive radiation oncology workflow. Recent work has begun to investigate SyntheticMR in the adaptive radiation oncology workflow, however no studies have investigated the variation in acquisition parameters and their relationship to the resulting quantitative maps. Filling this gap will facilitate SyntheticMR’s translation to the adaptive radiation therapy setting in the head and neck due to the reduced bias and increase repeatability and reproducibility of its generated quantitative relaxometric biomarkers.

**Approach:** The 2D multi-echo multi-delay (MDME) sequence offered from SyntheticMR was acquired on a MR-Simulation and MR-Linac device in both a phantom and healthy volunteers. Scans were optimized across acceleration factors, slice gaps, acquired voxel sizes, repetition times, echo times, refocusing flip angles, noise-limited gradients, number of slices, and echo train lengths. Quantitative relaxometric T1, T2, and PD maps were tested for accuracy using both correlation and mean absolute bias analysis. Noise profiles were evaluated using the coefficient of variation (CoV) in uniform regions of interest.

**Main Results:** The following main findings were reported: (1) noise-limited gradients did not affect the bias or CoV, (2) increasing acceleration factor did not affect bias, but it significantly increased CoV, (3) increasing the number of slices resulted in different significant changes in the quantitative parameters between the MR-Simulator and MR-Linac, (4) reducing the echo train length led to generally reduced bias and CoV, and (5) the repeatability CoV was within previously reported literature values on similar scanners.

**Significance:** The behavior of the 2D-MDME sequence from SyntheticMR was characterized across a wide range of acquisition parameters designed for ideal head and neck imaging on both the MR-Simulation and MR-Linac devices in phantom and healthy volunteers. Significant differences existed across several acquisition parameters which should be accounted for when making adjustments for application-specific field-of-views, scan time, and desired ranges of T1, T2, and PD. Application of these findings to the development of efficient head and neck adaptive radiation therapy MRI protocols can enrich the current quantitative biomarker landscape allowing for more informed treatment adaptations.

## 1. Introduction

As the accessibility of magnetic resonance imaging (MRI) and its wide range of capabilities expands^1^, its advantages over traditional imaging techniques such as computed tomography (CT) grow, particularly in the field of radiation oncology where optimal contrast is desired for lesion visualization^2^. For this reason, MRI has been steadily incorporated into the daily workflow of the radiation oncology department through MRI-based simulation scanners^3^ and MRI-guided radiation therapy delivery devices such as the MR-Linac^4^. These devices especially require consistent measurements over time to monitor for radiation induced changes which may trigger a treatment plan adaptation. Therefore, moving away from intensity-based changes (i.e., hypo-intense vs. hyper-intense) and towards quantitative measurements will enable optimal treatment adaptation protocols^5^. The basic measures of quantitative MRI-based tissue properties are T1, T2, and proton density (PD)^6^. In head and neck cancer specifically, these fundamental quantitative metrics have been studied as a means of diagnosing the initial lesion^7^, assessing its response to radiation and/or systemic therapies^8^, and how the surrounding normal tissue may be unintentionally damaged as a result^9^. However, these mapping techniques have traditionally been time consuming, thus limiting their clinical implementation^6,10,11^.

Overcoming this hurdle, the company SyntheticMR AB (Linköping, Sweden) has developed a method for simultaneous multiparametric MRI relaxometry quantification (T1, T2, and PD) in a single sequence originally known as QRAPMASTER^12^ and more recently adopted into industry as Multi-Dynamic Multi-Echo (MDME) on Siemens MRI scanners, MAGnetic resonance image Compilation (MAGiC) on GE MRI scanners, and Synthetic Acquisition (SyntAc) on Philips MRI scanners. Through their post-processing software, SyMRI, these acquired images can be reconstructed into synthetic contrast maps such as traditional contrast weighted images as well as customizable inversion recovery (IR) pre-pulses to nullify fluid (FLAIR), fat (STIR), and correct for phase (PSIR). Since all images derive from a single acquisition, as a result, every reconstructed image is inherently co-registered allowing for rapid comparison across tissue types. The most common, 2D multi-slice version of this sequence (2D-MDME)^13–15^, can acquire in-plane voxel sizes within 2 mm while sacrificing through-plane resolution to 3 – 6 mm depending on a preferred signal-to-noise ratio (SNR) for the desired anatomical site^16^.

Currently, literature regarding the optimization of the 2D-MDME sequence is limited. A previous paper has investigated adjusting the repetition time (TR) and echo time (TE) on the synthetically generated images from SyMRI for the shoulder through radiologist interpretation and found that shorter TE and longer TR improved contrast^17^. Another study asked neuroradiologists to optimize the TR, TE, and effective inversion time (TI_eff_) in SyMRI to achieve the ideal contrast to visualize the subthalamic nucleus^18^. However, limited work has been done to optimize the quantitative maps from SyntheticMR prior to processing in SyMRI. One abstract from the International Society of Magnetic Resonance in Medicine (ISMRM) annual meeting focused on the precision and accuracy of the 2D-MDME sequence in phantoms and the human brain^19^.

In radiation therapy, 2D-MDME has been shown to provide sufficient accuracy, repeatability, and reproducibility for treatment planning in the prostate^20^. Similarly, in the head and neck region for radiation therapy, a recent work explored the technical feasibility of integrating SyntheticMR 2D-MDME acquisitions into the head and neck cancer adaptive radiation oncology workflow^21^, demonstrating clinically acceptable quantification of T1, T2, and PD while maintaining a logistically feasible acquisition time in under six minutes. This work provides a framework to expand on previous studies such as one finding that demyelination in normal appearing white matter in glioma patients as a result of radiation therapy can be detected using quantitative MRI^22^. Though the previously stated studies are promising, unanswered questions remained such as: Can the 2D-MDME sequence provide more accurate quantification than that presented in the study? Can the total acquisition time be reduced to streamline the clinical workflow without hindering this accuracy? How stable are subtle changes in acquisition parameters on the resulting quantitative maps? Therefore, in this paper, we plan to expand upon the previously stated work by using the radiotherapy-predicate studies, idea, development, exploration, assessment, and long-term study (R-IDEAL) framework, as recommended by the MR-Linac Consortium, to complete a Stage 2a (technical optimization) systematic evaluation^23^. The expected outcome of this study is to identify optimal acquisition parameters based on clinical requirements while maintaining clinically feasible acquisition times and safe delivery.

## 2. Methods and Materials

### 2.1. MRI Acquisition Parameters

To evaluate SyntheticMR across the radiation oncology department, MRI scans were performed on a 3T MR-Simulator scanner (XA50 / XA60, Vida; Siemens Healthcare; Erlangen, Germany) and a 1.5T MR-Linac (R5.7.1.2, Unity; Elekta AB; Stockholm, Sweden). For this study, the 2D-MDME sequence was acquired across several optimization dimensions as shown in **Table 1**.

**Table 1.** MRI acquisition parameter optimization workflow for both the MR-Simulator and MR-Linac. Note that some features are available on the MR-Linac which are not for the MR-Simulator such as adjusting the refocusing flip angle and adding noise reduction techniques. Additionally, since the MR-Simulator is Siemens and the MR-Linac is Philips, they both present with different acceleration methods (GRAPPA^24^ for Siemens and CS-SENSE^25^ for Philips).

|  | MR-Simulator | MR-Linac |
| --- | --- | --- |
| → Step 1 | Slice Thickness<br>Slice Gap | Slice Thickness<br>Slice Gap<br>Refocusing Flip Angle<br>SofTone Acoustic Noise Reduction |
| → Step 2 | GRAPPA Factor | CS-SENSE Factor<br>Denoising Factor |
| → Step 3 | Echo Time (TE) | Echo Time (TE) |
| → Step 4 | Number of Slices<br>Repetition Time (TR) | Number of Slices<br>Repetition Time (TR) |
| → Step 5 | Turbo Factor | TSE Factor |
| → Step 6 | Final Repeatability Assessment | Final Repeatability Assessment |

**Step 1.** The slice thickness was varied between 3 mm and 4 mm for the MR-Simulator and just 3 mm for the MR-Linac. Similarly, the slice gap was kept at 10% for the MR-Simulator (i.e., 0.3 mm and 0.4 mm, respectively) while switching between 10% (0.3 mm) and 33% (1 mm) for the MR-Linac. A slice gap of at least 10% is recommended by SyntheticMR AB to reduce cross-talk which may result in overestimated T2 quantification^12^. Further, a minimum slice thickness of 3 mm is recommended to maintain adequate SNR. The refocusing flip angle on the MR-Simulator could only be set to 150° so it was not optimized for that scanner. This parameter could be adjusted on the MR-Linac and was set to either 120° or 160°. The MR-Linac also had SofTone acoustic noise reduction available which was tested due to the loud nature of the MDME sequence. Our hypothesis was that the increased slice thickness would increase the signal-to-noise ratio (SNR) while compromising quantification due to partial volume effects, increased slice gap would improve quantification due to reduced slice cross-talk, larger refocusing flip angles would increase the SNR, and that SofTone would reduce SNR.

**Step 2.** For scan acceleration purposes, the MR-Simulator had GRAPPA available which was modified between factors of 1 to 5. Secondly, the MR-Linac had CS-SENSE available which was modified between factors of 2 to 3 due to the increased time constraints while maintaining sufficient SNR and stable g-factors at increasing acceleration factors. Thirdly, the MR-Linac also had varying denoising factors available for the CS-SENSE reconstruction which was tested for both weak and strong strengths. Our hypothesis was that increased acceleration factors should simultaneously degrade quantification and noise performance.

**Step 3.** A total of six different echo time combinations for the MR-Simulator (9.4 / 94, 19 / 94, 28 / 94, 9.4 / 113, 19 / 113, and 28 / 113 ms) and nine different echo time combinations for the MR-Linac (11 / 107, 21 / 108, 12 / 117, 22 / 117, 31 / 118, 13 / 127, 24 / 127, and 33 / 127 ms) were tested. Note that the equivalent of 28 / 94 ms on the MR-Simulator could not be performed in the MR-Linac due to gradient limitations. Our hypothesis was that these echo time combinations should primarily affect the T2 quantification.

**Step 4.** The number of slices for both the MR-Simulator and MR-Linac were increased between 30 and 60 which also correspondingly increased the repetition time (TR) and acquisition time.

Our hypothesis was that the TR should primarily affect the T1 quantification due to the accommodated TI_eff_. Note that the inversion time is referred to as “effective” here due to the actual inversion time staying constant between dynamics, but the images acquired for T1 quantification are created by shifting the frequency of the radiofrequency (RF)-inversion pulse relative to the frequency of the slice RF-excitation pulse.

**Step 5.** Since we decided to use the fast spin echo version of SyntheticMR’s MDME sequence instead of GRaSE^26^, we could adjust the echo train length via a turbo factor of 5 and 6 for the MR-Simulator and TSE factor of 10 and 12 for the MR-Linac. Both of these adjustments lead to an echo train length of 10 and 12 since the 2D-MDME sequence natively employs two echo times per dynamic. Our hypothesis was that reduced echo train length would positively affect quantification and SNR due to the increased signal from fewer echoes compared to lower signal from echoes further down the echo train^27^.

**Step 6.** Repeatability was assessed by repeating the determined best acquisition from Steps 1 – 5 a total of five times successively in the same scan session.

All healthy volunteer scans were acquired in the axial / transverse orientation to best visualize structures in the head and neck^28,29^ while phantom scans were acquired coronally. Note that a 200 mm inferior-superior coverage at the in-plane field-of-view (FOV) of 256 x 256 mm^2^ was decided as required to adequately visualize the head and neck region in the superior-inferior extent. A fixed pixel bandwidth of 210 Hz/pixel and 231 Hz/pixel was used for the MR-Simulator and MR-Linac, respectively.

### 2.2. MRI Phantoms Assessed

The CaliberMRI “ISMRM/NIST” Premium System Phantom Model 130 phantom (CaliberMRI; Boulder, CO) was used as a reference for NIST-traceable T1, T2, and PD values^30^. This phantom includes 14 vials for each metric suitable for T1 values between 20 and 1724 ms, T2 values between 9 and 853 ms, and PD values between 5 and 100% at 20°C on a 1.5T MRI scanner. Reference values are also provided by the CaliberMRI for 3T MRI scanners. The bore temperature was monitored using a manual thermometer which regularly read between 20°C and 21°C across all acquisitions.

### 2.3. Healthy Volunteer Description

One healthy male and one healthy female volunteer were imaged on the MR-Simulator using the determined best sequence to evaluate clinical reproducibility. One healthy male volunteer was imaged on the MR-Linac for step 1 of the optimization schedule from **Table 1**.

### 2.4. Image Processing and Analysis

The images were processed using the SyntheticMR post-processing software, SyMRI (StandAlone 11.3.11; SyntheticMR AB; Linköping, Sweden) developed specifically for the 2D-MDME sequence. For the phantom analysis, a circular region-of-interest (ROI) was created in each vial using 3D Slicer^31^ (https://www.slicer.org/) using the quantitative T1 map (SYMAP) and the values within each ROI were extracted for processing in the quantitative T1, T2, and PD maps. A margin of approximately 10% of the vial’s diameter was left to account for possible ringing artifacts^32^ and partial volume effects at the edges^33^. For the *in vivo* analysis, the left and right parotid and submandibular glands were chosen for analysis due to their critical role in salivary dysfunction throughout radiation therapy^34^ and were contoured automatically using a deep learning algorithm in the Advanced Medical Imaging Research Engine (ADMIRE) research software (v3.48.4; Elekta AB; Stockholm, Sweden). These automatic contours were validated by experienced medical physicians (Z.B., C.D., W.F., A.M.S.A., A.S.R.M.) in RayStation Research 12A R v13.1.100.0 (RaySearch Laboratories; Stockholm, Sweden).

### 2.5. Statistical Analysis

All relevant analysis concerning statistical methods were formulated using the guidelines for reporting Statistical Analyses and Methods in the Published Literature (SAMPL)^35^. All computational analysis was completed using Python 3.8.10 and is freely available at https://github.com/Lucas-Mc/SyntheticMR_R-IDEAL_2a. To evaluate quantitative parameter accuracy, for each vial inside the ISMRM/NIST phantom, the mean values inside the ROI were calculated and compared to CaliberMRI, NIST-traceable, reference values. The standard deviation values of each vial we also extracted to evaluate varying noise behavior across quantitative parameters. The mean absolute bias instead of the mean bias was used for evaluation due to its un-signed behavior, thus reducing errors where equal positive and negative bias would result in a mean bias of 0%. Lin’s Concordance Correlation Coefficient^36^ (LCCC) was used to evaluate direct agreement to reference values instead of generalized linearity while the coefficient of variation (CoV) was used as a measure of relative quantification variation and noise. Further, we chose to limit our analysis to clinical T1 values in the range of 250 – 2000 ms, T2 values in the range of 30 – 200 ms, and PD values in the range of 20 – 160 pu^37^ for both the 3T MR-Simulator and 1.5T MR-Linac. A p-value of 0.05 was used for statistical significance.

For the MR-Simulator, the Wilcoxon signed-rank test^38^ was used to evaluate the difference between the mean absolute bias for the varying slice thickness and turbo factor acquisitions. The Kruskal-Wallis test^39^ was then used to evaluate equal acquisitions across varying acceleration factors. In the healthy volunteers, cluster analysis was performed using an elliptical envelope and a soft margin assuming 25% outliers while resulting statistical significance testing between each healthy volunteer scan across combined laterality salivary glands was calculated using the SigClust^40,41^ method with soft thresholding and 1,000 Monte Carlo iterations following mean centering and variance normalization. For the MR-Linac, the Wilcoxon signed-rank test and Hedges’ g effect size^42^ was used to evaluate the difference between the mean absolute bias for the varying slice gap, CS-SENSE factor, denoising factor, refocusing flip angle, SofTone acoustic noise reduction factor, and TSE factor acquisitions. For both the MR-Simulator and MR-Linac, the Pearson correlation coefficient^43^ and subsequent linear line of best fit was used to compare trends in mean absolute bias across increasing number of slices. This was also done on the MR-Simulator across increasing acceleration factors. Further, repeatability was quantified by using the LCCC and mean absolute bias for the quantitative T1, T2, and PD values and calculating the CoV across the repeated acquisitions for each metric. The optimal echo time acquisition was determined by searching for the lowest combined mean absolute bias while giving preference to T1 and T2 since this is used most often in clinical settings for head and neck cancer^9,44^.

## 3. Results

### 3.1. MR-Simulator Phantom Analysis

**Steps 1 – 3.** For a slice thickness of 3 mm and slice gap of 0.3 mm, adjusting the GRAPPA acceleration factor from 1 to 5 resulted in a scan time reduction from 13:28 to 3:31. In most cases, a GRAPPA factor of 4 minimized the T1 mean absolute bias while a GRAPPA factor of 3 minimized most T2 mean absolute bias cases. For PD mean absolute bias, a GRAPPA factor of 1 provided the minimum result most cases. Looking across the six different echo time combinations, little difference (around 1 – 2%) was seen across the T1 mean absolute bias, however large differences are seen in both the T2 and PD mean absolute bias. The T2 mean absolute bias was minimized generally at echo times of 28 / 113 ms while the PD mean absolute bias was minimized at echo times of 9.4 / 94 ms. However, to determine the best combination across T1, T2, and PD, the cumulative mean absolute bias must be taken into account which is minimized for echo times of 19 / 94 ms. Therefore, this combination of echo times was chosen when conducting future optimizations beginning after Step 4.

Analyzing the same experimental setup after changing the slice thickness from 3 mm to 4 mm and the slice gap from 0.3 mm to 0.4 mm yielded the following results. Adjusting the GRAPPA acceleration factor from 1 to 5 resulted in the same scan time reduction from 13:28 to 3:31. In most cases, a GRAPPA factor of 4 still minimized the T1 mean absolute bias while a GRAPPA factor of 1 minimized most T2 mean absolute bias cases. For PD mean absolute bias, a GRAPPA factor of 5 minimized most cases, however its difference was minimal at 1 – 2%.

Statistically, there were no significant changes as shown in **Figure 1**. A similar analysis was done for the change in the maximum CoV which found a significant increase in PD (r = 0.95) in a slice thickness of 3 mm and both T1 (r = 0.95) and PD (r = 0.91) in a slice thickness of 4 mm also shown in **Figure 1**

**Figure 1.**
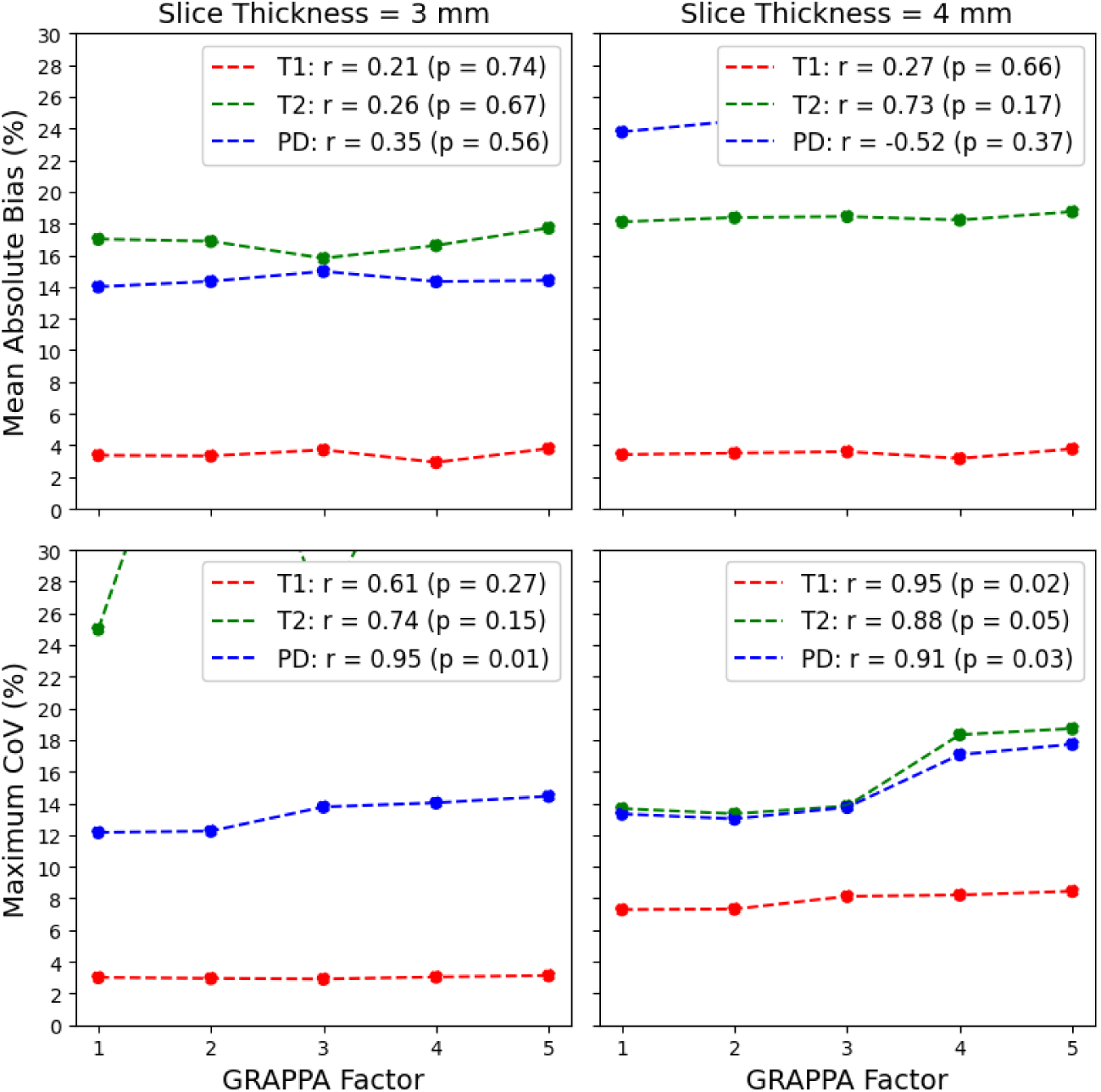
Top: Average change in the quantitative T1, T2, and PD mean absolute bias on the MR-Simulator as the GRAPPA factor increases. Bottom: Average change in the quantitative T1, T2, and PD maximum CoV on the MR-Simulator as the GRAPPA factor increases. Note, a maximum CoV of 30% was set to show the trends since the PD maximum CoV of 95% for a GRAPPA factor of 5.

Looking across the six different echo time combinations (9.4 / 94, 19 / 94, 28 / 94, 9.4 / 113, 19 / 113, and 28 / 113 ms), little difference (around 1 – 2%) is seen across the T1 mean absolute bias again, however large differences are seen in both the T2 and PD mean absolute bias. The T2 mean absolute bias rose from 2 – 5% to 40% when the first echo time was set to 9.4 ms while the PD mean absolute bias rose from 16 – 18% to 30% when the first echo was larger than 9.4 ms. As with a slice thickness of 3 mm and a slice gap of 0.3 mm, the T2 mean absolute bias was minimized generally at echo times of 28 / 113 ms while the PD mean absolute bias was minimized at echo times of 9.4 / 113 ms. The same best echo times of 19 / 94 ms held for this setup when the slice thickness was 4 mm and the slice gap was 0.4 mm, however, overall the mean absolute bias increased across the T1, T2, and PD values indicating that a 3 mm slice thickness and 0.3 mm slice gap should be used moving forward. Statistically, the one-sided Wilcoxon signed-rank test showed a p-value of 0.19 for the T1 mean absolute bias (greater than), <0.0001 for T1 (less than), and <0.0001 for PD (less than) from 3 mm slice thickness / 0.3 mm slice gap to 4 mm slice thickness / 0.4 mm slice gap. Therefore, the smaller slice thickness and slice gap of 3 mm and 0.3 mm, respectively, provide both a more favorable mean absolute bias while also delivering increased spatial resolution. The Kruskal-Wallis test across increasing GRAPPA factors for the 3 mm slice thickness / 0.3 mm slice gap combination showed significant difference among the T1 mean absolute bias (p = 0.03) but not for the T2 mean absolute bias (p = 0.88), and PD mean absolute bias (p = 0.95). For the T1 mean absolute bias, the GRAPPA factor that showed a minimum was 4, however, we decided to continue with a GRAPPA factor of 3 to limit potential acceleration-induced artifacts *in vivo*. More detailed results are shown in **Figure 2**, **Figure S1**, and **Figure S2**.

**Figure 2.**
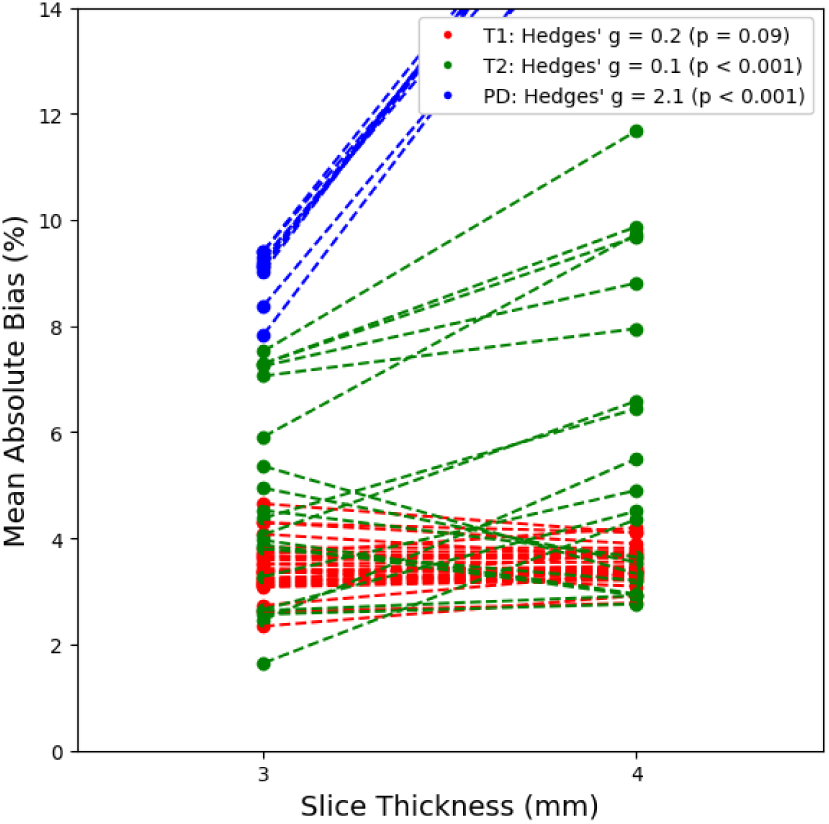
Change in quantitative T1, T2, and PD mean absolute bias on the MR-Sim as the slice thickness increases from 3 mm to 4 mm. Note, a maximum mean absolute bias of 14% was set to only include the clinically feasible sequences.

**Step 4.** Varying the number of slices from 30 to 60 resulted in a scan time range between 3:19 to 6:39 with every slice adding around 6 – 7 seconds of added scan time despite the variations in the TR. Interestingly, the first TI_eff_ stayed constant at 173 ms across all acquisitions. The second TI_eff_ increased by around 146 ms every 10 slices (i.e., >30, >40, etc.). The third TI_eff_ increased by around 75 ms every slice, however sometimes it stayed constant despite a 2 slice increase. The fourth and final TI_eff_ increased by around 85 ms every slice. The final TI_eff_ remained 120 ms below the TR for each acquisition. Increasing the number of slices had negligible effect on the T1 and T2 mean absolute bias but increased PD mean absolute bias. As a result, every reduction in around 10 slices (or 33 mm in coverage with a 3 mm slice thickness and 0.3 mm slice gap) results in an approximate 1 – 2% reduction in PD mean absolute bias. This appears primarily driven by the changing second TI_eff_ every 10 slices. Although the minimum PD mean absolute bias was achieved by minimizing the TR, and thus, the number of slices, we decided to choose 50 slices as our chosen best clinical sequence for the MR-Simulator due to its increased coverage, allowing us to characterize more normal tissue properties throughout the course of radiation therapy. The Pearson correlation coefficient testing of mean absolute bias against increasing number of slices was r =-0.76 (p = 0.0007) for T1, r = 0.52 (p = 0.04) for T2, and r = 0.96 (p < 0.0001) for PD demonstrating increased bias with respect to increasing number of slices. However, the line of best fit was *-0.01*n_slices + 3.98* for T1, *0.02*n_slices + 3.26* for T2, and *0.10*n_slices + 8.34* for PD showing the slowest increasing bias for T1 followed by T2 then PD. Even in the case of PD, the bias is only expected to increase by 1% every 10 slices while every 50 slices for T2 and every 100 slices for T1 showcasing sufficient stability for most dynamic field-of-view examinations. More detailed results are shown in **Figure 3** and **Figure S3**.

**Figure 3.**
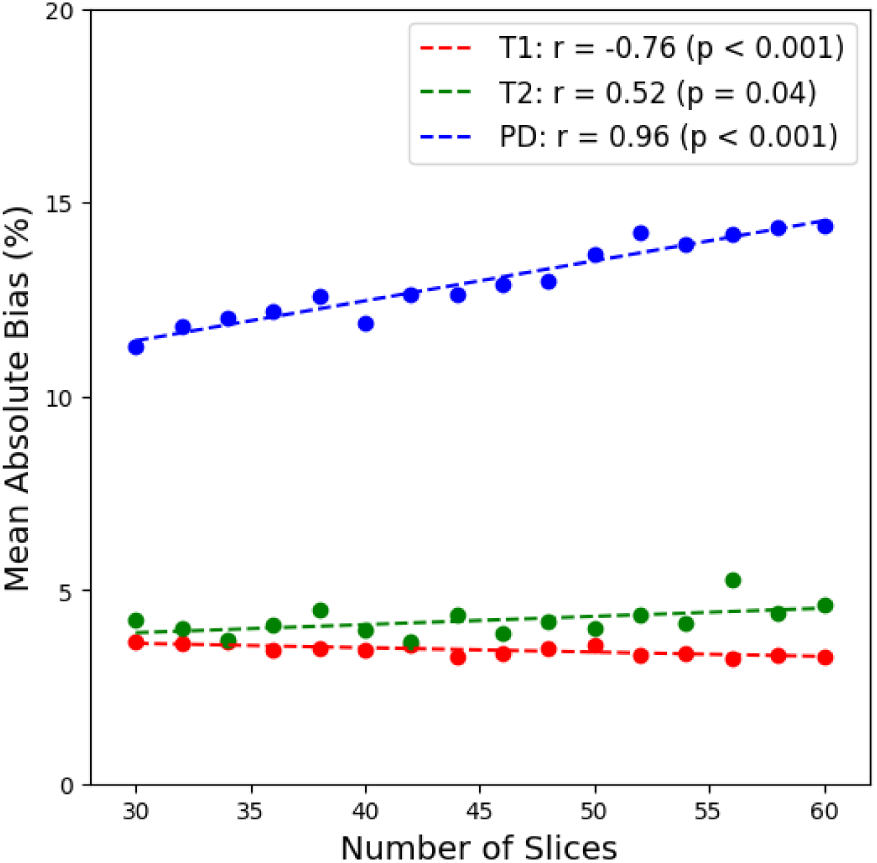
Change in quantitative T1, T2, and PD mean absolute bias on the MR-Simulator as the number of slices increases.

**Step 5.** Only turbo factors of 5 and 6 were available for this set of acquisition parameters. Reducing the turbo factor to 5 from 6 which was used for all previous acquisitions lead to a reduced mean absolute bias for T1 (3.47 % to 2.52%), T2 (4.37 % to 3.36%), and PD (13.65% to 11.65%) at the compromise of an extra minute of scan time (5:31 to 6:29). Although the reduced bias is beneficial, its magnitude was not large enough to sacrifice the extra minute of scan time for our MR simulation protocol which already was at maximum allowed scan time.

Understanding that these acquisitions will be acquired repeatedly for the purpose of anatomical changes instead of diagnostic accuracy further influenced this opinion and led to our next experiment of quantifying repeatability. More detailed results are shown in **Figure S4**.

**Step 6.** The repeatability as measured by the CoV across five repetitions of the LCCC values was less than 0.1% for T1, T2, and PD. Further, the CoV across five repetitions of the mean absolute bias was within 3% for T1, within 7% for T2, and within 2% for PD. More detailed results are shown in **Figure S5**.

### 3.2. MR-Simulator Healthy Volunteer Analysis

Visually, as shown in **Figure 4**, the elliptical envelopes for the same healthy volunteer across repetitions are nearly identical across all cluster analyses. The largest difference appeared to be in the T1 vs. PD cluster. Volunteer 2 appeared to have generally the same quantitative values as Volunteer 1 with some discrepancies, primarily across T1 and PD with very similar results across T2.

**Figure 4.**
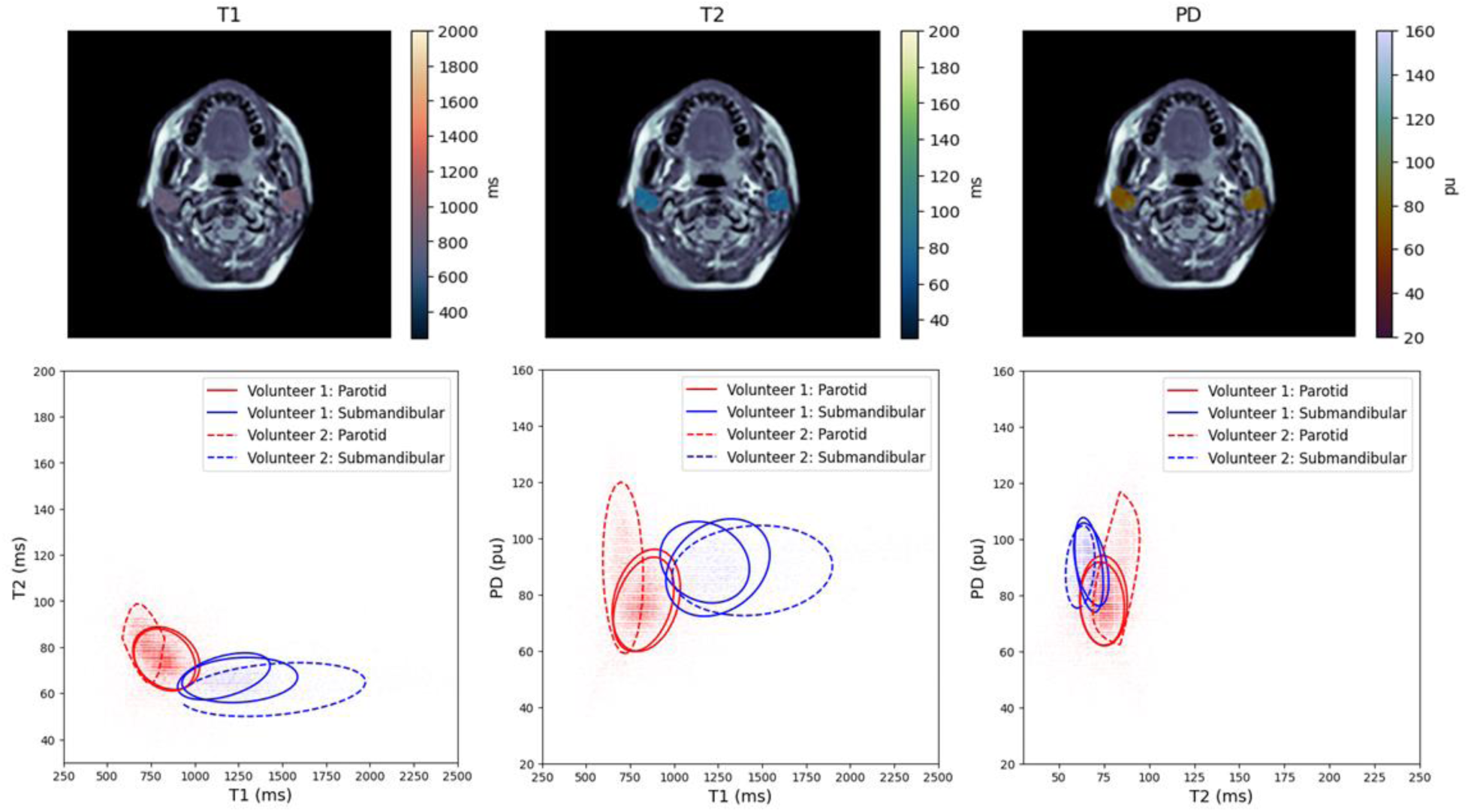
Top: Sample anatomical image slice from a healthy volunteer with the overlayed SyntheticMR generated quantitative T1 (left), T2 (middle), and PD (right) maps. Bottom: A cluster analysis in the combined parotid and submandibular salivary glands when comparing quantitative T1 and T2 (left), T1 and PD (middle), and T2 and PD (right). Note, Volunteer 1 had two repeated scans which is shown as two of the same styled ellipses.

Statistically, all p-values from SigClust were greater than 0.86 indicating no statistical significance between any clusters from the same type of salivary gland across all healthy volunteer scans. Although the T1 in the submandibular gland in Volunteer 2 was larger than in Volunteer 1 (1604 ms vs. ∼1220 ms), the standard deviation in the T1 value for Volunteer 2 was 455 ms leading to significant cluster overlap, thus removing possible statistical significance between clusters. Similarly, the T1 in the parotid gland in Volunteer 2 was smaller than in Volunteer 1 (736 ms vs. ∼850 ms) though the standard deviation was 175 ms in Volunteer 2 leading to no statistical significance between the clusters. All PD values were similar across all volunteers and salivary gland. A tabular summary of the statistical results is shown in **Table 2**.

**Table 2.** Mean and standard deviation (std) values for the three health volunteer scans across the quantitative T1, T2, and PD clusters.

| | **T1 (ms)**<br>mean ± std | **T2 (ms)**<br>mean ± std | **PD (pu)**<br>mean ± std |
| --- | --- | --- | --- |
| **Volunteer 1 / Rep. 1** |  |  |  |
| Parotid | 854 ± 143 | 75 ± 9 | 78 ± 11 |
| Submandibular | 1212 ± 252 | 69 ± 9 | 91 ± 8 |
| **Volunteer 1 / Rep. 2** |  |  |  |
| Parotid | 849 ± 132 | 73 ± 9 | 77 ± 9 |
| Submandibular | 1233 ± 226 | 66 ± 6 | 90 ± 9 |
| **Volunteer 2** |  |  |  |
| Parotid | 736 ± 175 | 81 ± 29 | 87 ± 17 |
| Submandibular | 1604 ± 455 | 63 ± 8 | 89 ± 8 |

### 3.3. MR-Linac Phantom Analysis

**Step 1 – 2.** The mean absolute bias changed significantly from a slice gap of 0.3 mm to 1 mm with a decrease in T1 (Hedges’ g =-1.3), increase in T2 (Hedges’ g = 0.4), and decrease in PD (Hedges’ g =-0.3). Switching the CS-SENSE factor from 2 to 3 lead to a significant increase in mean absolute bias in T1 (Hedges’ g = 1.1), T2 (Hedges’ g = 1.4), and PD (Hedges’ g = 0.2).

The strong denoising factor slightly increased the mean absolute bias significantly for T1 (Hedges’ g = 0.3) and T2 (Hedges’ g = 0.2) with no significant change for PD. Strong effects were seen in the mean absolute bias when increasing the refocusing flip angle from 120° to 160° with an increase for T1 (Hedges’ g = 1.1), decrease in T2 (Hedges’ g =-2.5), and decrease in PD (Hedges’ g =-6.9). No significant changes in the mean absolute bias were seen in any quantitative parameters (T1, T2, and PD) when enabling to SofTone. More detailed results are shown in **Figure 5**, **Figure S6**, and **Figure S7**.

**Figure 5.**
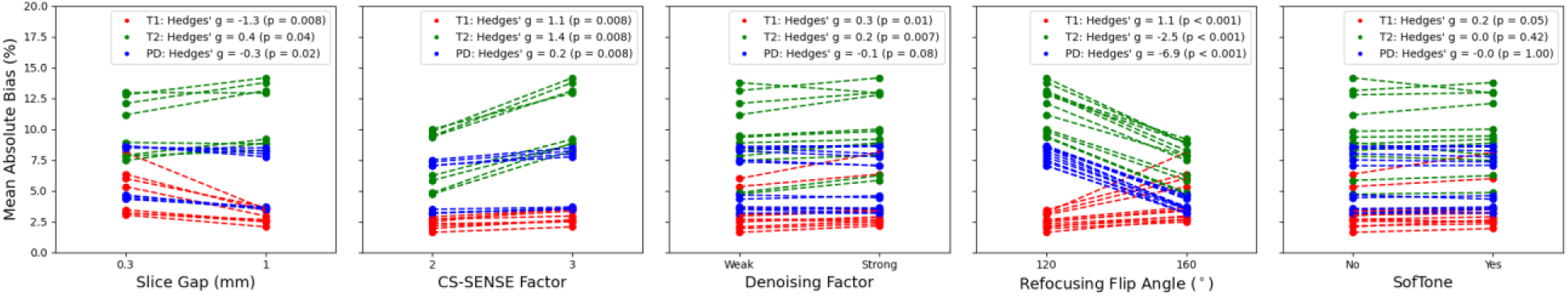
Change in quantitative T1, T2, and PD mean absolute bias on the MR-Linac across varying slice gaps, CS-SENSE factors, denoising factor, refocusing flip angles, and SofTone acoustic noise reduction.

Figure 5. Change in quantitative T1, T2, and PD mean absolute bias on the MR-Linac across varying slice gaps, CS-SENSE factors, denoising factor, refocusing flip angles, and SofTone acoustic noise reduction.

**Step 3.** For the echo time optimization, the minimum T2 mean absolute bias for each set of second echo times (i.e., 108, 117, 127 ms) occurred at the middle first echo time (i.e., 21, 22, 24 ms, respectively). Conversely, the minimum PD mean absolute bias for each set of second echo times (i.e., 107, 117, 127 ms) occurred at the smallest first echo time (i.e., 11, 12, 13 ms, respectively). Interestingly, the T1 mean absolute bias remained relatively constant across the varying echo time combinations. Therefore, when combining this evidence together, we decided to continue exploring the 24 / 127 ms echo time combination for the number of slices and TSE factor optimization. More detailed results are shown in **Figure S8**.

**Step 4.** When comparing previously chosen echo times of 24 / 127 ms across acquisitions with 30 – 50 slices, the TR increased from 5551 ms to 9252 ms approximately by 185 ms per added slice. The resulting acquisition time difference ranged from 3:48 to 6:19 increasing by around 7.5 seconds per slice. Interestingly, the first TI_eff_ stayed the same even when the number of slices increased. The second TI_eff_ increased by 185 ms every 10 slices while the third TI_eff_ increased by approximately 185 ms every two slices, however this trend was not consistent. The fourth TI_eff_ increased by 185 ms every slice. The Pearson correlation coefficient testing of mean absolute bias against increasing number of slices was r =-0.15 (p = 0.67) for T1, r =-0.84 (p = 0.0013) for T2, and r = 0.40 (p = 0.22) for PD demonstrating significant decreased bias with respect to increasing number of slices for T2 quantification. However, the line of best fit was *-0.08*n_slices + 8.98* for T2 demonstrating that the bias is only expected to increase by 1% every 13 slices showcasing sufficient stability for most dynamic field-of-view examinations. The resulting FOV can be determined by *n_slices * (slice_thickness + slice gap) – slice_gap*. Therefore, 50 slices with a slice thickness of 3 mm and slice gap of 1 mm will cover a range of *50 * (3 mm + 1 mm) – 1 mm = 199 mm*. This coverage is desired for head and neck cancer due to the wide range of anatomical locations the tumor may reside and the large spread of normal tissues affected ranging from the esophagus to the brain. Therefore, moving forward, we decided to choose the 50 slice acquisition due to its large coverage (199 mm). This paired with a decreasing T2 mean absolute bias compared to an acquisition with fewer slices while maintaining the same T1 and PD mean absolute bias was a critical motivating factor for pursuing a higher number of slices. More detailed results are shown in Figure 6 and **Figure S9**.

**Figure 6.**
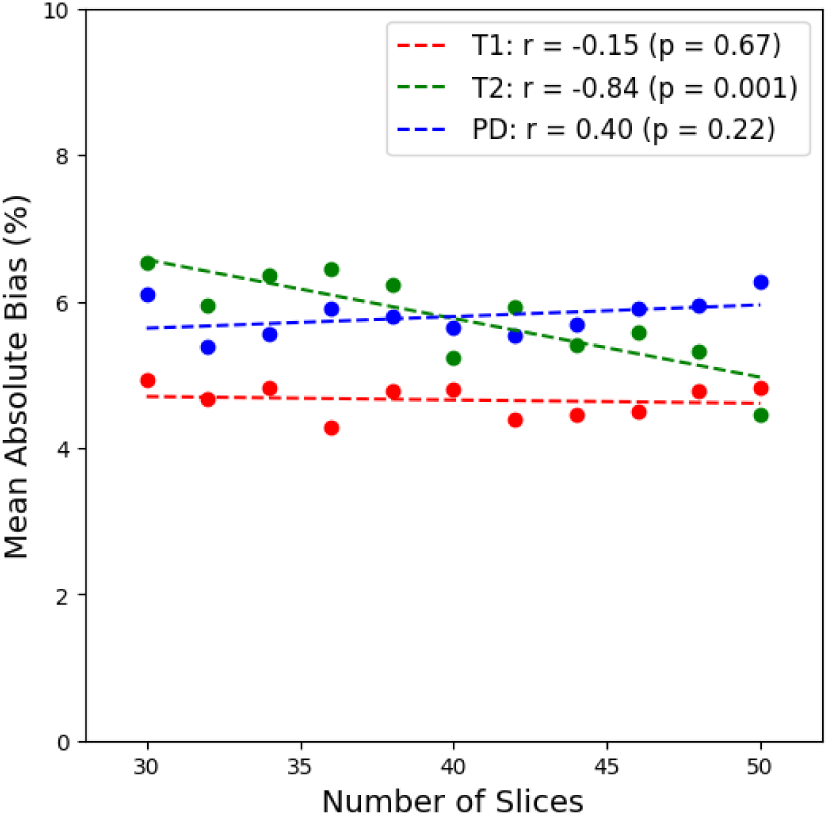
Change in quantitative T1, T2, and PD mean absolute bias on the MR-Linac as the number of slices increases.

**Step 5.** Only TSE factors of 12 and 14 were feasible at the desired scan time limitations and echo time combination. Setting the TSE factor to 10 would have results in a scan time over 7 minutes 30 seconds which was above our clinically feasible range. Unlike the MR-Simulator which showed a reduced mean absolute bias at lower TSE / turbo factors across the quantitative metrics, the MR-Linac, at our chosen acquisition settings only showed reduced mean absolute bias for T2 quantification (5.32% vs. 3.60%). Interestingly, the T1 mean absolute bias increased (4.80% vs. 5.59%) while the PD mean absolute bias was similar (6.54% vs. 6.68%). All LCCC values were above 0.98. From these results, we chose to use the TSE factor of 12 due to its reduced T1 mean absolute bias and perceived reduced overall image noise due to the reduced loss of signal in the echoes at the end of the echo train in 12 vs. 14 total echoes. More detailed results are shown in **Figure S10**.

**Step 6.** The CoV across five consecutive repeats of the final chosen sequence was less than 0.3% across T1, T2, and PD when analyzing the direct agreement between measured quantitative values and phantom reference values through the LCCC. Additionally, the CoV across repeated mean absolute bias measurements was within 5% for T1, 6% for T2, and 2% for PD. More detailed results are shown in **Figure S11**.

### 3.4. MR-Linac Healthy Volunteer Analysis

The graphical representation of the CoV results are shown in Figure 7 for each of the salivary glands (left parotid gland, right parotid gland, left submandibular gland, and right submandibular gland) and quantitative parameters (T1, T2, and PD). In summary, a slice gap of 1 mm consistently and significantly (p < 0.05) reduced the CoV compared to a slice gap of 0.3 mm, particularly in the quantitative T1 values by an average Hedges’ g effect size of-1.6 across all salivary glands. On the other hand, the average effect size was-0.6 for T2 and-1.3 for PD with neither of the submandibular glands providing significant changes from the Wilcoxon signed-rank test for each. When comparing the different CS-SENSE factors, limited statistical significance was seen, however in the four times that it occurred (T2 in the left parotid, T1 in the right parotid, T1 in the right submandibular, and T2 in the right submandibular), each subgroup showed increased CoV in the CS-SENSE factor of 3 group except for the T2 in the left parotid where the CoV was higher in the CS-SENSE factor of 2 group. When comparing weak denoising to strong denoising, the effect size for the CoV change was negative across all quantitative parameters (T1, T2, and PD) for every region of interest (left parotid, right parotid, left submandibular, and right submandibular) indicating reduced noise for the strong denoising setting, as expected. The reduction was the strongest amongst the T2 maps with an average Hedges’ g effect size of-1.1 followed by PD then T1 with an average Hedges’ g effect size of - 0.6 and-0.55, respectively. However, the PD reduction was only significant in the left submandibular glands. Interestingly, no metric had a significant reduction in the right submandibular gland. Additionally, the T1 reduction was not significant in the right submandibular gland. The refocusing flip angle of 160° compared to 120° resulted in only three significant changes with increases in the CoV in the quantitative T1 (Hedges’ g = 0.7) and PD (Hedges’ g = 1.0) in the left parotid gland and T1 (Hedges’ g = 0.9) in the left submandibular gland. No significant changes were found in the CoV in any structure or quantitative value between SofTone being enabled or disabled.

**Figure 7.**
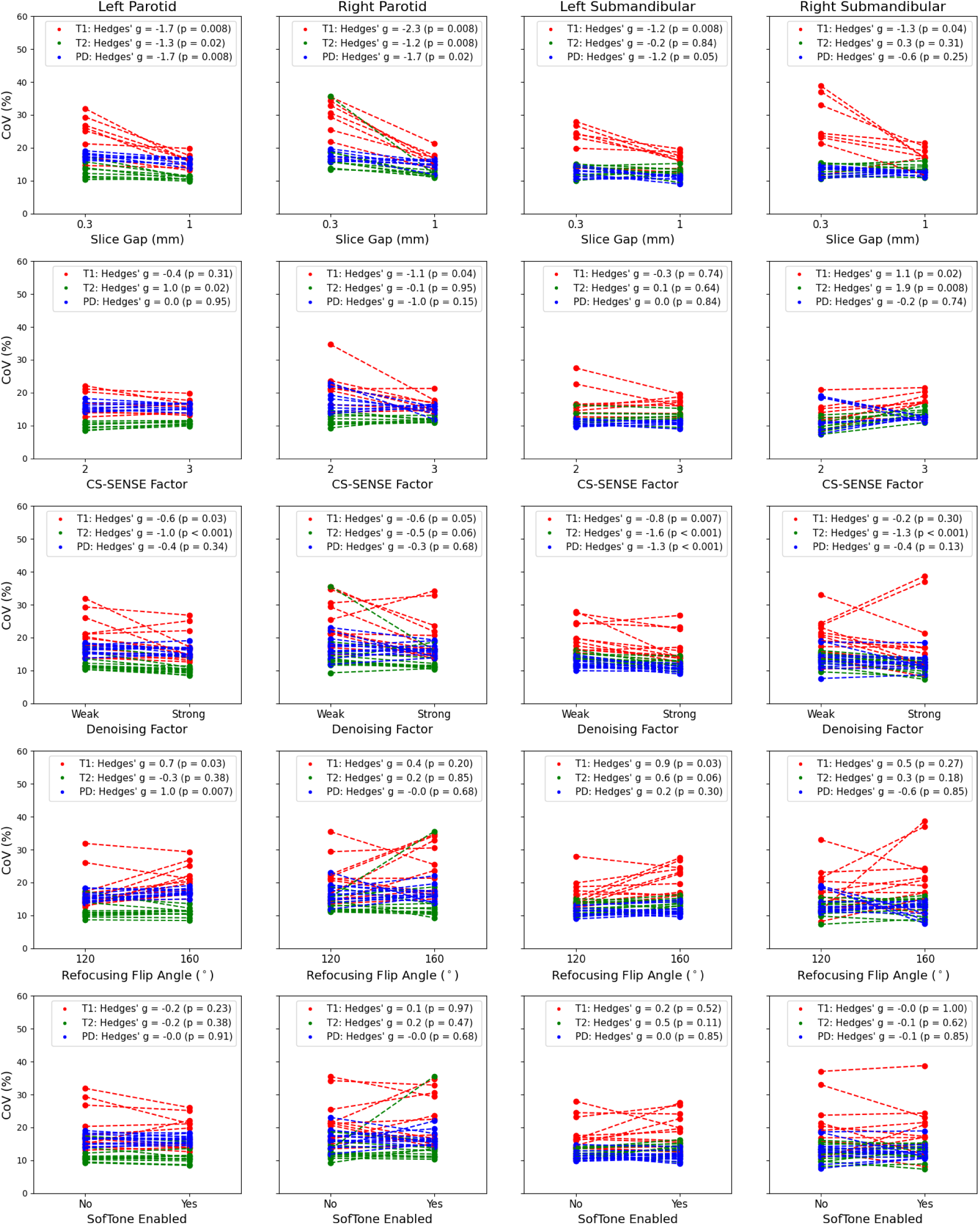
Change in quantitative T1, T2, and PD CoV on the MR-Linac in a healthy volunteer’s left parotid (far left), right parotid (center left), let submandibular (center right), and right submandibular (far right) salivary glands across varying slice gaps, CS-SENSE factors, denoising factor, refocusing flip angles, and SofTone acoustic noise reduction.

## 4. Discussion

In summary, we found that significant differences existed between the 2D-MDME acquisition parameters that affected the mean absolute bias and CoV amongst the quantitative T1, T2, and PD values calculated from the sequence. First, contrary to our predictions, the CoV did not change in any structure or quantitative value with SofTone enabled or disabled^45^, implying no change in the noise profile in presumed to be uniform structures. Additionally, the mean absolute bias did not change significantly between SofTone being disabled or enabled implying it to be a suitable alternative for protocols with existing high specific absorption rate^46^ (SAR) or in higher risk patients. Second, the GRAPPA factor did not have a consistent effect on the mean absolute bias, however the maximum CoV increased significantly in PD for both slice thicknesses of 3 mm and 4 mm while increasing in T1 for a slice thickness of 4 mm as the GRAPPA factor increases. Third, SyntheticMR AB, in their user manual, recommends a TR of 4000 ms to provide TI_eff_ sufficient for characterization of T1 values in most clinically relevant tissues. We saw that gradually increasing the TR through increasing the number of slices results in decreasing T1 mean absolute bias, increasing T2 mean absolute bias, and increasing PD mean absolute bias in the MR-Simulator. However, on the MR-Linac, the T2 mean absolute bias decreased with increasing TR while remaining the same for T1 and PD. There may be several factors involved here including static magnetic field strength’s influence on T1 values^47^, reduced slice gap for the MR-Simulator, and difference in acceleration techniques possibly influencing the reconstructed images. We did not show whether this effect holds when the number of slices is kept the same and the TR is adjusted manually away from its scanner provided default value. Another recommendation is to limit the coverage to no more than 50 slices, possibly due to the increased TR which may provide TI_eff_ too large for lower T1 tissues.

However, we saw that the T1 characterization as measured by the mean absolute bias stayed relatively constant across varying TRs. Moreso, surprisingly, the TI_eff_ adjusted with periodic frequency depending on their order (i.e., first echo time stayed the same, the second echo time changed every 10 slices, etc.) instead of adjusting each TI_eff_ by a ratio in accordance with the changing TR. Knowledge of this information is useful when adjusting sequence parameters on the fly without time to perform quality assurance. Fourth, the repeatability CoV for the T1 and T2 mean absolute bias was within previously reported literature values on similar scanners^48,49^ while the PD mean absolute bias was slightly higher than previously reported^50^.

The recommended slice thickness for the MDME sequence is between 3 – 6 mm with main tradeoffs of SNR. For example, changing the slice thickness from 4 mm to 3 mm has a 4/3 = 1.33 reduction in SNR. For the best visualization of tumors and healthy structures in the head and neck area, we decided to commit to a 3 mm slice thickness despite its lower SNR since it still provided better quantitative maps than our current protocol. Further, it offered the lowest mean absolute bias across T1, T2, and PD measurements on the MR-Simulator. Although slice gap is recommended to be between 0.1 – 1 mm, we decided on a minimum slice gap of 10% for the MR-Simulator to reduce possible slice crosstalk effects while maintaining higher resolution in the slice orientation as compared to a typical slice gap of 1 mm. However, we decided on a slice gap of 1 mm for the MR-Linac due to the reduced SNR from the lower static magnetic field strength and weaker gradient system. This reduced gradient performance on the MR-Linac is why the ideal echo time of 19/94 ms on the MR-Simulator could not be achieved. Although the first echo time of 19 ms could be achieved, the gradients were not fast enough to acquire the second echo in less than 107 ms. Although our chosen CS-SENSE factor with strong denoising could potentially smooth small structures, our purpose of developing this SyntheticMR sequence was to monitor quantitative properties of larger normal structures throughout the course of radiation therapy. Therefore, we chose the strong denoising factor not only for its minimal effect on mean absolute bias, but also is ability to remove noise through a lower CoV in healthy volunteers which may hinder accurate quantification^51^.

Potential limitations of this study include not acquiring all investigated acquisition parameters and instead, following the workflow shown in **Table 1**. Therefore, potentially more optimal sequence exist where our data was not acquired. Further, scans were only conducted in healthy volunteers for a small amount of the chosen optimizations for the sequence, therefore limiting our full understanding of how these acquisition parameter changes affect quantitative metrics inside structures of interest. Another limitation of this study is the lack of investigations into optimization acquisition parameters such as bandwidth, TR above the minimum value, and FOV.

Recently, a 3D version of the 2D-MDME sequence capable of 1 mm isotropic acquisitions (3D-QALAS)^52^ has been developed and has shown quantitative accuracy and repeatability within clinical recommendations in both a multi-center^53^ and multi-vendor study^54^. A patch for this sequence exists for Philips on version R5.7.1.2 and we plan to investigate its feasibility using the MR-Linac’s native anterior / posterior coil combination within acceptable acquisition times in the future. The design of this sequence is fundamentally different than the 2D-MDME sequence which will require separate optimizations; however, it remains unknown if the same general trends will hold. In agreement with the R-IDEAL Stage 2a framework, comprehensive results have been provided here to aid in 2D-MDME protocol optimizations in other anatomical sites besides the head and neck. For example, knowledge of the tradeoffs between the number of slices and the respective quantitative maps on a relative scale can help to optimize protocols without exhaustive searches such as the one provided here.

## 5. Conclusion

This study investigated the technical optimization of the 2D-MDME sequence provided by SyntheticMR and its derivative quantitative T1, T2, and PD values in the head and neck. Scans were acquired on a MR-Simulation and MR-Linac device in both a phantom and healthy volunteers where they were optimized across acceleration factors, slice gaps, acquired voxel sizes, repetition times, echo times, refocusing flip angles, and noise-limited gradients.

Significant differences existed across several acquisition parameters which should be accounted for when making adjustments for application-specific field-of-views, scan time, and desired ranges of T1, T2, and PD.

## Data Availability

All relevant anonymized SyntheticMR quantitative maps and contours in NIfTI format are to be made publicly available after manuscript acceptance at the following DOI: 10.6084/m9.figshare.28703528. The accompanying code for image visualization and statistical analysis will be made publicly available at the following URL: https://github.com/Lucas-Mc/SyntheticMR_R-IDEAL_2a.

https://doi.org/10.6084/m9.figshare.28703528

https://github.com/Lucas-Mc/SyntheticMR_R-IDEAL_2a

## Acknowledgements

The authors would like to thank SyntheticMR AB for their assistance in the formal data analysis through their software, SyMRI.

