## Supplementary Materials for "Technical Optimization of SyntheticMR for the Head and Neck on a 3T MR-Simulator and 1.5T MR-Linac: A Prospective R-IDEAL Stage 2a Technology Innovation Report"

### MR-Simulator:

**Table S1.** Echo time and acceleration factor optimization acquired with 50 slices using slice thickness = 3 mm, slice gap = 0.3 mm, TR = 7310 ms, and TI = 173 / 1050 / 3389 / 7190 ms.

| GRAPPA Factor | $TE_1$ / $TE_2$ (ms) | Acq. Time (mm:ss) | T1 Mean Absolute Bias (%) | T2 Mean Absolute Bias (%) | PD Mean Absolute Bias (%) |
|---|---|---|---|---|---|
| 1 | 9.4 / 94 | 13:28 | 3.37 | 43.21 | 9.12 |
| 2 | 9.4 / 94 | 7:21 | 3.21 | 43.85 | 9.03 |
| 3 | 9.4 / 94 | 4:57 | 3.83 | 42.91 | 9.15 |
| 4 | 9.4 / 94 | 3:59 | 3.35 | 41.12 | 9.41 |
| 5 | 9.4 / 94 | 3:31 | 3.70 | 43.69 | 7.84 |
| 1 | 19 / 94 | 13:28 | 3.60 | 7.06 | 14.75 |
| 2 | 19 / 94 | 7:21 | 3.74 | 4.07 | 16.10 |
| 3 | 19 / 94 | 4:57 | 4.30 | 2.46 | 17.59 |
| 4 | 19 / 94 | 3:59 | 3.26 | 4.38 | 15.90 |
| 5 | 19 / 94 | 3:31 | 4.65 | 2.66 | 16.52 |
| 1 | 28 / 94 | 13:28 | 3.13 | 3.88 | 18.30 |
| 2 | 28 / 94 | 7:21 | 3.10 | 4.53 | 18.57 |
| 3 | 28 / 94 | 4:57 | 3.17 | 3.28 | 18.58 |
| 4 | 28 / 94 | 3:59 | 2.34 | 5.36 | 18.29 |
| 5 | 28 / 94 | 3:31 | 3.52 | 4.95 | 19.50 |
| 1 | 9.4 / 113 | 13:28 | 3.39 | 38.15 | 9.41 |
| 2 | 9.4 / 113 | 7:21 | 3.11 | 38.91 | 9.21 |
| 3 | 9.4 / 113 | 4:57 | 3.52 | 38.63 | 9.28 |
| 4 | 9.4 / 113 | 3:59 | 2.74 | 37.58 | 9.14 |
| 5 | 9.4 / 113 | 3:31 | 3.19 | 43.71 | 8.38 |
| 1 | 19 / 113 | 13:28 | 3.65 | 7.25 | 14.85 |
| 2 | 19 / 113 | 7:21 | 3.66 | 7.29 | 15.40 |
| 3 | 19 / 113 | 4:57 | 4.08 | 5.91 | 16.89 |
| 4 | 19 / 113 | 3:59 | 3.24 | 7.29 | 15.23 |
| 5 | 19 / 113 | 3:31 | 4.29 | 7.53 | 15.72 |
| 1 | 28 / 113 | 13:28 | 3.08 | 2.57 | 17.62 |
| 2 | 28 / 113 | 7:21 | 3.15 | 2.64 | 17.80 |
| 3 | 28 / 113 | 4:57 | 3.39 | 1.65 | 18.39 |
| 4 | 28 / 113 | 3:59 | 2.61 | 3.97 | 18.02 |
| 5 | 28 / 113 | 3:31 | 3.43 | 3.80 | 18.48 |

**Table S2.** Echo time and acceleration factor optimization acquired with 50 slices using slice thickness = 4 mm, slice gap = 0.4 mm, TR = 7210 ms, and TI = 169 / 1034 / 3341 / 7090 ms.

| GRAPPA Factor | $TE_1$ / $TE_2$ (ms) | Acq. Time (mm:ss) | T1 Mean Absolute Bias (%) | T2 Mean Absolute Bias (%) | PD Mean Absolute Bias (%) |
|---|---|---|---|---|---|
| 1 | 9.4 / 94 | 13:28 | 3.48 | 45.93 | 17.96 |
| 2 | 9.4 / 94 | 7:21 | 3.71 | 46.18 | 18.33 |
| 3 | 9.4 / 94 | 4:57 | 3.82 | 44.13 | 17.95 |
| 4 | 9.4 / 94 | 3:59 | 3.39 | 45.32 | 18.25 |
| 5 | 9.4 / 94 | 3:31 | 4.20 | 44.95 | 17.10 |
| 1 | 19 / 94 | 13:28 | 3.63 | 7.95 | 24.24 |
| 2 | 19 / 94 | 7:21 | 3.76 | 6.59 | 26.19 |
| 3 | 19 / 94 | 4:57 | 3.90 | 5.50 | 25.08 |
| 4 | 19 / 94 | 3:59 | 3.37 | 6.44 | 24.72 |
| 5 | 19 / 94 | 3:31 | 4.11 | 4.51 | 25.30 |
| 1 | 28 / 94 | 13:28 | 3.39 | 2.92 | 29.48 |
| 2 | 28 / 94 | 7:21 | 3.28 | 3.65 | 30.08 |
| 3 | 28 / 94 | 4:57 | 3.44 | 4.91 | 29.34 |
| 4 | 28 / 94 | 3:59 | 2.92 | 3.33 | 29.99 |
| 5 | 28 / 94 | 3:31 | 3.56 | 3.55 | 28.76 |
| 1 | 9.4 / 113 | 13:28 | 3.10 | 40.27 | 17.69 |
| 2 | 9.4 / 113 | 7:21 | 3.27 | 41.25 | 18.03 |
| 3 | 9.4 / 113 | 4:57 | 3.37 | 42.04 | 17.45 |
| 4 | 9.4 / 113 | 3:59 | 3.35 | 41.41 | 18.13 |
| 5 | 9.4 / 113 | 3:31 | 3.38 | 44.61 | 16.73 |
| 1 | 19 / 113 | 13:28 | 3.66 | 8.81 | 24.58 |
| 2 | 19 / 113 | 7:21 | 3.71 | 9.69 | 24.83 |
| 3 | 19 / 113 | 4:57 | 3.64 | 9.73 | 24.37 |
| 4 | 19 / 113 | 3:59 | 3.21 | 9.87 | 24.25 |
| 5 | 19 / 113 | 3:31 | 4.11 | 11.68 | 24.18 |
| 1 | 28 / 113 | 13:28 | 3.23 | 2.76 | 28.76 |
| 2 | 28 / 113 | 7:21 | 3.34 | 2.92 | 29.36 |
| 3 | 28 / 113 | 4:57 | 3.44 | 4.36 | 28.73 |
| 4 | 28 / 113 | 3:59 | 2.78 | 2.95 | 28.91 |
| 5 | 28 / 113 | 3:31 | 3.31 | 3.20 | 27.73 |

**Table S3.** Number of slices and TR optimization using slice thickness = 3 mm, slice gap = 0.3 mm, GRAPPA factor = 3, and TE = 19 / 94 ms.

| TR (ms) | Slices | $TI_1$ / $TI_2$ / $TI_3$ / $TI_4$ (ms) | Acq. Time (mm:ss) | T1 Mean Absolute Bias (%) | T2 Mean Absolute Bias (%) | PD Mean Absolute Bias (%) |
|---|---|---|---|---|---|---|
| 4390 | 30 | 173 / 612 / 2075 / 4270 | 3:19 | 3.65 | 4.24 | 11.27 |
| 4680 | 32 | 173 / 758 / 2220 / 4560 | 3:32 | 3.62 | 3.99 | 11.82 |
| 4970 | 34 | 173 / 757 / 2219 / 4850 | 3:45 | 3.65 | 3.72 | 12.04 |
| 5270 | 36 | 173 / 758 / 2369 / 5150 | 3:59 | 3.47 | 4.10 | 12.21 |
| 5560 | 38 | 173 / 758 / 2514 / 5440 | 4:12 | 3.52 | 4.48 | 12.59 |
| 5850 | 40 | 173 / 758 / 2659 / 5730 | 4:25 | 3.45 | 3.96 | 11.89 |
| 6140 | 42 | 173 / 904 / 2804 / 6020 | 4:38 | 3.59 | 3.68 | 12.64 |
| 6430 | 44 | 173 / 903 / 2949 / 6310 | 4:51 | 3.29 | 4.37 | 12.65 |
| 6730 | 46 | 173 / 904 / 3099 / 6610 | 5:04 | 3.37 | 3.89 | 12.87 |
| 7020 | 48 | 173 / 904 / 3244 / 6900 | 5:18 | 3.51 | 4.18 | 12.96 |
| 7310 | 50 | 173 / 1050 / 3389 / 7190 | 5:31 | 3.60 | 4.00 | 13.67 |
| 7600 | 52 | 173 / 1050 / 3388 / 7480 | 5:44 | 3.23 | 4.36 | 14.22 |
| 7900 | 54 | 173 / 1051 / 3538 / 7780 | 5:58 | 3.34 | 4.15 | 13.91 |
| 8190 | 56 | 173 / 1050 / 3863 / 8070 | 6:11 | 3.24 | 5.25 | 14.17 |
| 8480 | 58 | 173 / 1050 / 3828 / 8360 | 6:24 | 3.34 | 4.40 | 14.34 |
| 8770 | 60 | 173 / 1196 / 3973 / 8650 | 6:37 | 3.30 | 4.63 | 14.42 |

**Table S4.** Turbo factor optimization using 50 slices with slice thickness = 3 mm, slice gap = 0.3 mm, GRAPPA factor = 3, TE = 19 / 94 ms, TR = 7310 ms, and TI = 173 / 1050 / 3389 / 7190 ms.

| Turbo Factor | Acq. Time (mm:ss) | T1 LCCC | T2 LCCC | PD LCCC | T1 Mean Absolute Bias (%) | T2 Mean Absolute Bias (%) | PD Mean Absolute Bias (%) |
|---|---|---|---|---|---|---|---|
| 5 | 6:29 | 0.9989 | 0.9965 | 0.9805 | 2.52 | 3.36 | 11.65 |
| 6 | 5:31 | 0.9974 | 0.9951 | 0.9733 | 3.47 | 4.37 | 13.65 |

**Table S5.** Repeatability analysis using 50 slices with slice thickness = 3 mm, slice gap = 0.3 mm, GRAPPA factor = 3, TE = 19 / 94 ms, TR = 7310 ms, TI = 173 / 1050 / 3389 / 7190 ms, turbo factor = 6, and acquisition time = 5:31.

| Repeat # | T1 LCCC | T2 LCCC | PD LCCC | T1 Mean Absolute Bias (%) | T2 Mean Absolute Bias (%) | PD Mean Absolute Bias (%) |
|---|---|---|---|---|---|---|
| **1** | 0.9976 | 0.9976 | 0.9732 | 3.43 | 4.17 | 13.91 |
| **2** | 0.9977 | 0.9965 | 0.9723 | 3.25 | 4.30 | 14.03 |
| **3** | 0.9977 | 0.9952 | 0.9732 | 3.24 | 4.75 | 13.93 |
| **4** | 0.9974 | 0.9957 | 0.9719 | 3.32 | 4.88 | 14.11 |
| **5** | 0.9974 | 0.9951 | 0.9733 | 3.47 | 4.37 | 13.65 |
| **CoV (%)** | 0.01 | 0.09 | 0.06 | 2.77 | 6.10 | 1.11 |

**MR-Linac:**

**Table S6.** Image quality optimization. Each acquisition had 60 slices with a slice thickness = 3 mm with a TE = 12 / 120 ms. Note that the slice gap of 0.3 mm with the desired superior-inferior coverage was not feasible at a CS-SENSE acceleration factor of 2. The field-of-view was 284 x 284 mm$^2$ with an acquired voxel size of 1.00 x 1.00 mm$^2$ reconstructed to 1.00 x 1.00 mm$^2$.

| Slice Gap (mm) | CS-SENSE Factor | Denoising Factor | Refocusing Flip Angle (°) | SofTone Enabled | TR (ms) | $TI_1$ / $TI_2$ / $TI_3$ / $TI_4$ (ms) | Acq. Time (mm:ss) |
|---|---|---|---|---|---|---|---|
| 1 | 2 | Weak | 120 | No | 8976 | 149 / 1196 / 4039 / 8826 | 6:09 |
| 1 | 2 | Weak | 120 | Yes | 8988 | 149 / 1198 / 4044 / 8838 | |
| 1 | 2 | Weak | 160 | No | 9001 | 150 / 1200 / 4050 / 8850 | |
| 1 | 2 | Weak | 160 | Yes | 9014 | 150 / 1201 / 4056 / 8863 | |
| 1 | 2 | Strong | 120 | No | 8976 | 149 / 1196 / 4039 / 8826 | |
| 1 | 2 | Strong | 120 | Yes | 8988 | 149 / 1198 / 4044 / 8838 | |
| 1 | 2 | Strong | 160 | No | 9001 | 150 / 1200 / 4050 / 8850 | |
| 1 | 2 | Strong | 160 | Yes | 9014 | 150 / 1201 / 4056 / 8863 | |
| 0.3 | 3 | Weak | 120 | No | 10772 | 179 / 1436 / 4847 / 10592 | 4:30 |
| 0.3 | 3 | Weak | 120 | Yes | 10788 | 179 / 1438 / 4854 / 10608 | |
| 0.3 | 3 | Weak | 160 | No | 10803 | 180 / 1440 / 4861 / 10622 | |
| 0.3 | 3 | Weak | 160 | Yes | 10819 | 180 / 1442 / 4868 / 10638 | |
| 0.3 | 3 | Strong | 120 | No | 10772 | 179 / 1436 / 4847 / 10592 | |
| 0.3 | 3 | Strong | 120 | Yes | 10788 | 179 / 1438 / 4854 / 10608 | |
| 0.3 | 3 | Strong | 160 | No | 10803 | 180 / 1440 / 4861 / 10622 | |
| 0.3 | 3 | Strong | 160 | Yes | 10819 | 180 / 1442 / 4868 / 10638 | |
| 1 | 3 | Weak | 120 | No | 8976 | 149 / 1196 / 4039 / 8826 | 3:45 |
| 1 | 3 | Weak | 120 | Yes | 8988 | 149 / 1198 / 4044 / 8838 | |
| 1 | 3 | Weak | 160 | No | 9001 | 150 / 1200 / 4050 / 8850 | |

| | | | | | | |
|---|---|---|---|---|---|---|
| 1 | 3 | Weak | 160 | Yes | 9014 | 150 / 1201 / 4056 / 8863 |
| 1 | 3 | Strong | 120 | No | 8976 | 149 / 1196 / 4039 / 8826 |
| 1 | 3 | Strong | 120 | Yes | 8988 | 149 / 1198 / 4044 / 8838 |
| 1 | 3 | Strong | 160 | No | 9001 | 150 / 1200 / 4050 / 8850 |
| 1 | 3 | Strong | 160 | Yes | 9014 | 150 / 1201 / 4056 / 8863 |

**Table S7.** Phantom image quality optimization. Each acquisition had 50 slices with a slice thickness = 3 mm with a TE = 12 / 120 ms. The field-of-view was 284 x 284 mm$^2$ with an acquired voxel size of 1.00 x 1.00 mm$^2$ reconstructed to 1.00 x 1.00 mm$^2$.

| Slice Gap (mm) | CS-SENSE Factor | Denoising Factor | Refocusing Flip Angle (°) | SofTone Enabled | T1 Mean Absolute Bias (%) | T2 Mean Absolute Bias (%) | PD Mean Absolute Bias (%) |
|---|---|---|---|---|---|---|---|
| 1 | 2 | Weak | 120 | No | 1.63 | 9.35 | 7.51 |
| 1 | 2 | Weak | 120 | Yes | 1.94 | 9.43 | 7.34 |
| 1 | 2 | Weak | 160 | No | 2.68 | 4.72 | 3.20 |
| 1 | 2 | Weak | 160 | Yes | 2.47 | 4.84 | 3.50 |
| 1 | 2 | Strong | 120 | No | 2.16 | 9.80 | 7.03 |
| 1 | 2 | Strong | 120 | Yes | 2.33 | 9.99 | 8.06 |
| 1 | 2 | Strong | 160 | No | 2.68 | 5.84 | 3.18 |
| 1 | 2 | Strong | 160 | Yes | 2.89 | 6.23 | 3.20 |
| 0.3 | 3 | Weak | 120 | No | 3.04 | 11.15 | 8.47 |
| 0.3 | 3 | Weak | 120 | Yes | 3.18 | 12.08 | 8.59 |
| 0.3 | 3 | Weak | 160 | No | 5.33 | 7.81 | 4.65 |
| 0.3 | 3 | Weak | 160 | Yes | 5.99 | 7.46 | 4.31 |
| 0.3 | 3 | Strong | 120 | No | 3.46 | 12.77 | 8.60 |
| 0.3 | 3 | Strong | 120 | Yes | 3.23 | 12.96 | 8.66 |
| 0.3 | 3 | Strong | 160 | No | 6.34 | 8.91 | 4.43 |
| 0.3 | 3 | Strong | 160 | Yes | 8.12 | 7.88 | 4.49 |
| 1 | 3 | Weak | 120 | No | 2.08 | 13.12 | 8.48 |
| 1 | 3 | Weak | 120 | Yes | 2.66 | 13.78 | 8.23 |
| 1 | 3 | Weak | 160 | No | 2.96 | 8.26 | 3.55 |
| 1 | 3 | Weak | 160 | Yes | 3.66 | 8.88 | 3.68 |
| 1 | 3 | Strong | 120 | No | 2.53 | 14.16 | 8.01 |
| 1 | 3 | Strong | 120 | Yes | 2.49 | 12.92 | 7.76 |
| 1 | 3 | Strong | 160 | No | 3.37 | 8.80 | 3.50 |
| 1 | 3 | Strong | 160 | Yes | 3.49 | 9.18 | 3.60 |

**Table S8.** Echo time optimization. The TR was set to the shortest possible value which is why it varies across acquisitions. Each acquisition had 50 slices with a slice thickness = 3 mm and slice gap = 1 mm, CS-SENSE factor = 2 with strong denoising, refocusing flip angle = 160°, TSE factor = 12, and no SofTone was applied. The field-of-view was 256 x 256 $mm^2$ with an acquired voxel size of 2.00 x 2.19 $mm^2$ reconstructed to 1.00 x 1.00 $mm^2$.

| $TE_1$ / $TE_2$ (ms) | TR (ms) | Acq. Time (mm:ss) | T1 Mean Absolute Bias (%) | T2 Mean Absolute Bias (%) | PD Mean Absolute Bias (%) |
|---|---|---|---|---|---|
| 11 / 107 | 8599 | 5:53 | 4.27 | 9.06 | 4.31 |
| 21 / 108 | 9199 | 6:17 | 4.14 | 5.67 | 6.97 |
| 12 / 117 | 8793 | 6:00 | 4.75 | 8.65 | 3.98 |
| 22 / 117 | 9199 | 6:17 | 4.53 | 4.93 | 6.46 |
| 31 / 118 | 9798 | 6:42 | 4.85 | 8.23 | 8.22 |
| 13 / 127 | 9487 | 6:29 | 4.90 | 7.83 | 3.97 |
| 24 / 127 | 9252 | 6:19 | 4.44 | 4.01 | 5.86 |
| 33 / 127 | 9798 | 6:42 | 4.15 | 8.58 | 8.29 |

**Table S9.** Repetition time optimization. Each acquisition had a slice thickness = 3 mm and slice gap = 1 mm, CS-SENSE factor = 2 with strong denoising, refocusing flip angle = 160°, TSE factor = 12, and no SofTone was applied. TE = 24 / 127 ms. The field-of-view was 256 x 256 $mm^2$ with an acquired voxel size of 2.00 x 2.19 $mm^2$ reconstructed to 1.00 x 1.00 $mm^2$. Note that the number of slices greater than 50 was not explored due to its excessive scan time compared to our desired 6-minute limitation.

| TR (ms) | Slices | $TI_1$ / $TI_2$ / $TI_3$ / $TI_4$ (ms) | Acq. Time (mm:ss) | T1 Mean Absolute Bias (%) | T2 Mean Absolute Bias (%) | PD Mean Absolute Bias (%) |
|---|---|---|---|---|---|---|
| 5551 | 30 | 185 / 740 / 2590 / 5365 | 3:48 | 4.93 | 6.54 | 6.10 |
| 5921 | 32 | 185 / 925 / 2775 / 5735 | 4:03 | 4.66 | 5.94 | 5.38 |
| 6291 | 34 | 185 / 925 / 2775 / 6105 | 4:18 | 4.81 | 6.36 | 5.56 |
| 6662 | 36 | 185 / 925 / 2960 / 6476 | 4:33 | 4.28 | 6.56 | 5.89 |
| 7032 | 38 | 185 / 925 / 3145 / 6846 | 4:48 | 4.77 | 6.23 | 5.80 |
| 7402 | 40 | 185 / 925 / 3330 / 7216 | 5:04 | 4.79 | 5.24 | 5.65 |
| 7772 | 42 | 185 / 1110 / 3515 / 7586 | 5:19 | 4.39 | 5.92 | 5.53 |
| 8142 | 44 | 185 / 1110 / 3700 / 7956 | 5:34 | 4.45 | 5.41 | 5.69 |
| 8512 | 46 | 185 / 1110 / 3885 / 8326 | 5:49 | 4.50 | 5.58 | 5.89 |
| 8882 | 48 | 185 / 1110 / 4070 / 8696 | 6:04 | 4.77 | 5.33 | 5.94 |
| 9252 | 50 | 185 / 1110 / 4255 / 9066 | 6:19 | 4.83 | 4.45 | 6.28 |

**Table S10.** TSE factor optimization. Each acquisition had a field-of-view was 256 x 256 $mm^2$ with an acquired voxel size of 2.00 x 2.19 mm reconstructed to 1.00 x 1.00 $mm^2$ with a slice thickness = 3 mm and slice gap = 1 mm, CS-SENSE factor = 2 with strong denoising, refocusing flip angle = 160°, and TE = 24 / 127 ms with no SofTone applied.

| TSE Factor | TR (ms) | Acq. Time (mm:ss) | T1 LCCC | T2 LCCC | PD LCCC | T1 Mean Absolute Bias (%) | T2 Mean Absolute Bias (%) | PD Mean Absolute Bias (%) |
|---|---|---|---|---|---|---|---|---|
| 12 | 9252 | 6:19 | 0.9864 | 0.9886 | 0.9946 | 4.80 | 5.32 | 6.54 |
| 14 | 10398 | 5:43 | 0.9819 | 0.9935 | 0.9940 | 5.59 | 3.60 | 6.68 |

**Table S11.** Each acquisition had a slice thickness = 3 mm and slice gap = 1 mm, CS-SENSE factor = 2 with strong denoising, refocusing flip angle = 160°, TSE factor = 12, and no SofTone was applied. TR = 9252 ms. TE = 24 / 127 ms. The field-of-view was 256 x 256 $mm^2$ with an acquired voxel size of 2.00 x 2.19 $mm^2$ reconstructed to 1.00 x 1.00 $mm^2$ in an acquisition time of 6:19.

| Repeat # | T1 LCCC | T2 LCCC | PD LCCC | T1 Mean Absolute Bias (%) | T2 Mean Absolute Bias (%) | PD Mean Absolute Bias (%) |
|---|---|---|---|---|---|---|
| **1** | 0.9876 | 0.9836 | 0.9947 | 4.37 | 4.88 | 6.32 |
| **2** | 0.9884 | 0.9792 | 0.9941 | 4.41 | 5.68 | 6.01 |
| **3** | 0.9868 | 0.9848 | 0.9948 | 4.86 | 5.43 | 6.18 |
| **4** | 0.9863 | 0.9847 | 0.9951 | 4.64 | 5.26 | 6.04 |
| **5** | 0.9889 | 0.9797 | 0.9949 | 4.23 | 5.56 | 6.20 |
| **CoV (%)** | 0.10 | 0.25 | 0.02 | 4.99 | 5.18 | 1.82 |
